# Optimising Cardiac Diffusion Tensor Imaging In Vivo: More Directions or Averages?

**DOI:** 10.1101/2025.02.08.25321885

**Authors:** Sam Coveney, David Shelley, Richard Foster, Maryam Afzali, Ana-Maria Poenar, Noor Sharrack, Sven Plein, Erica Dall’Armellina, Jürgen E. Schneider, Christopher Nguyen, Irvin Teh

## Abstract

**Background:** Cardiac diffusion tensor imaging (cDTI) is sensitive to imaging parameters including the number of unique diffusion encoding directions (ND) and number of repetitions (NR; analogous to number of signal averages or NSA). However, there is no clear guidance for optimising these parameters in the clinical setting.

**Methods:** Spin echo cDTI data with 2^nd^ order motion compensated diffusion encoding gradients were acquired in ten healthy volunteers on a 3T MRI scanner with different diffusion encoding schemes in pseudo-randomised order. The data were subsampled to yield 96 acquisition schemes with 6 ≤ ND ≤ 30 and 33 ≤ total number of acquisitions (NA_all_) ≤ 180. Stratified bootstrapping with robust fitting was performed to assess the precision and accuracy of each acquisition scheme. This was quantified across a mid-ventricular short-axis slice in terms of root mean squared difference (RMSD) with respect to the full reference dataset, and standard deviation (SD) across bootstrap samples respectively.

**Results:** For the same acquisition time, the ND = 30 schemes had on average 48%, 40%, 34% and 34% lower RMSD and 6.2%, 7.4%, 10% and 5.6% lower SD in MD, FA, HA and E2A compared to the ND = 6 schemes. Given a fixed number of high b-value acquisitions, there was a trend towards lower RMSD and SD of MD and FA with increasing numbers of low b-value acquisitions. Higher NA_all_ with longer acquisition times led to improved accuracy in all metrics whereby quadrupling NA_all_ from 40 to 160 volumes led to a 21%, 40%, 13% and 4.5% reduction in RMSD of MD, FA, HA and E2A, averaged across six diffusion encoding schemes. Precision was also improved with a corresponding 53%, 50%, 52% and 36% reduction in SD.

**Conclusions:** We observed that accuracy and precision were enhanced by (i) prioritising number of diffusion encoding directions over number of repetitions given a fixed acquisition time, (ii) acquiring sufficient low b-value data, (iii) using longer protocols where feasible. For clinically relevant protocols, our findings support the use of ND = 30 and NA_b50_:NA_b500_ ≥ 1/3 for better accuracy and precision in cDTI parameters. These findings are intended to help guide protocol optimisation for harmonisation of cDTI.

## INTRODUCTION

Cardiac diffusion tensor imaging (cDTI) is a rapidly emerging technique for myocardial tissue characterisation *in vivo* without need for contrast agents^1^. It has shown promise in characterising the microstructural changes in several clinical scenarios, including myocardial infarction^2–6^, hypertrophic cardiomyopathy^7–11^, aortic stenosis^12^, amyloidosis^13^ and others. The major challenge of cDTI for *in vivo* imaging is its high sensitivity to bulk motion of the heart during contracture. To ameliorate this, the use of 2^nd^ order motion-compensated (M2) diffusion encoding gradients has become standard practice in spin echo echo planar imaging (EPI)^14,15^. This leads to longer echo times (TEs) and lower signal-to-noise-ratio (SNR) compared to using non-motion-compensated diffusion gradients, which is compounded by the relatively short T_2_ of the myocardium, i.e. ∼44 ms at 3T^16^.

To obtain adequate SNR and to minimise artefacts, clinical cDTI protocols typically employ high numbers of repetitions (8 ≤ NR ≤ 16)^14,17,18^. We refer to number of repetitions (NR) instead of the more commonly used number of signal averages (NSA) because cardiac DTI data are in general exported and reconstructed offline as separate repetitions instead of being averaged on the scanner. However, the number of unique diffusion encoding directions (6 ≤ ND ≤ 12)^14,15,17–19^ often remains small relative to DTI protocols used in other anatomy such as the brain, where ND ≥ 30 has been recommended^20^. As the imaging time is related to both NR and ND, identifying a suitable range of NR and ND is important for cDTI within a clinically feasible timeframe. Moreover, DTI parameters, including mean diffusivity (MD) and fractional anisotropy (FA), as measured in the healthy myocardium with spin echo methods are known to vary in the literature with reports of 0.75 × 10^-3^ mm^2^/s ≤ MD ≤ 1.72 × 10^-3^ mm^2^/s and 0.29 ≤ FA ≤ 0.43 ^17,19,21^. Known imaging sources of variation for these parameters include spatial resolution, ND, NR, SNR, diffusion time and b-value. Isolating the effects of NR and ND is therefore an important aspect in the optimisation of efficient and robust cDTI protocols for clinical use.

In DTI, ND = 6 is the minimum number of unique high b-value acquisitions required for tensor estimation^22^. However, previous work in the brain investigating the dependence of DTI parameters on ND, showed that higher ND reduced artefactually elevated FA^23,24^ and its standard deviation (SD)^23^, particularly in regions of low FA^25^. Higher SNR, as may be obtained by averaging, led to similarly lower and more accurate FA^26,27^. As a guide, ND ≥ 20 was recommended for measurement of FA^23,28,29^ while ND ≥ 30 was suggested for measurement of tensor orientation and MD^23^. Whilst the effects of ND and NR have been reported ex vivo^30^, there have only been preliminary reports^31–34^ that provide no conclusive evidence on the requirements of ND and NR for cDTI *in vivo*. A recent consensus statement on cDTI published by the Society for Cardiovascular Magnetic Resonance Cardiac Diffusion Special Interest Group recommended that more than 6 directions should be used but highlighted that rationalising ND and NR remains an unmet need in the development of cDTI^1^.

In this study, we investigated the accuracy and precision of several diffusion sampling schemes over a wide range of ND and NR in healthy volunteers, extending our previous e*x vivo* work^30^. We compare time-normalised data to assess the trade-offs made between ND and NR, and examine the effects of the number of low b-value acquisitions. We hypothesized that accuracy and precision in DTI measurements would be improved by (i) sampling schemes that prioritise ND over NR, (ii) sufficient sampling of low b-value data, and (iii) longer scan times. This is intended to inform optimisation and standardisation of robust clinical cDTI protocols.

## METHODS

### Data Acquisition

Cardiac DTI data were acquired in healthy volunteers (N = 10) using a Prisma 3T MRI (Siemens Healthineers, Erlangen, Germany). The study was performed under approved ethics, and healthy volunteers provided written informed consent. Data were acquired with single-shot spin echo EPI, 2D radiofrequency inner volume excitation and cardiac triggering: TR = 3 RR-intervals, TE = 76 ms, in-plane resolution = 2.3 x 2.3 mm^2^, slice thickness = 8 mm, number of slices = 3, field-of-view = 320 x 111 mm^2^, partial Fourier = 7/8, bandwidth = 2012 Hz/px, b_low_ = 50 and b_high_ = 500 s/mm^2^. Up to 2^nd^ order motion compensated diffusion encoding gradient waveforms were applied. Subjects were scanned under free-breathing conditions without respiratory gating, in late systolic phase.

Diffusion-weighted data were acquired in pseudo-randomised fashion over a range of diffusion encoding schemes with different ND and NR. Low b-value data were acquired with ND = 3 orthogonal directions, denoted by ND_orth3, b50_. High b-value data were acquired with a 61-direction Cook diffusion encoding scheme^35^ that was sequentially subsampled to 6, 10, 18 and 30 direction sets. These diffusion encoding schemes (DES) were denoted Cook61_6, Cook61_10, Cook61_18 and Cook61_30. The Cook diffusion encoding scheme was particularly amenable for subsampling because it was optimised for incremental sampling in case of premature scan termination. Previous work has shown that noise performance (and therefore the accuracy of parameter estimates) of a given diffusion encoding scheme is significantly correlated with the condition number of its transformation matrix^36^. Specific sets of sequentially subsampled directions were identified to minimise the condition numbers of the transformation matrices. This was performed by circularly subsampling ND = 6, 10, 18 and 30 sequential directions from the Cook61 scheme, incrementing the first diffusion encoding direction (D_i_) from 1 to 61, and finding D_i_ that minimised the condition numbers. For the diffusion encoding schemes subsampled to 6, 10, 18 and 30 directions, condition numbers were relatively low at 1.74, 1.75, 1.70 and 1.58 respectively. For reference, the condition numbers of the widely used Jones30 and 6-direction dual gradient schemes are 1.59 and 2.00 respectively^36^. An icosahedral diffusion encoding scheme^37^ with 6 diffusion encoding directions (Icosa6) and Jones diffusion encoding scheme^38^ with 30 diffusion encoding directions (Jones30) were also acquired. An upper limit of 30 diffusion encoding directions was specified due to diminishing returns with higher ND^26^. These diffusion encoding schemes are illustrated in Figure 1.

**Figure 1.**
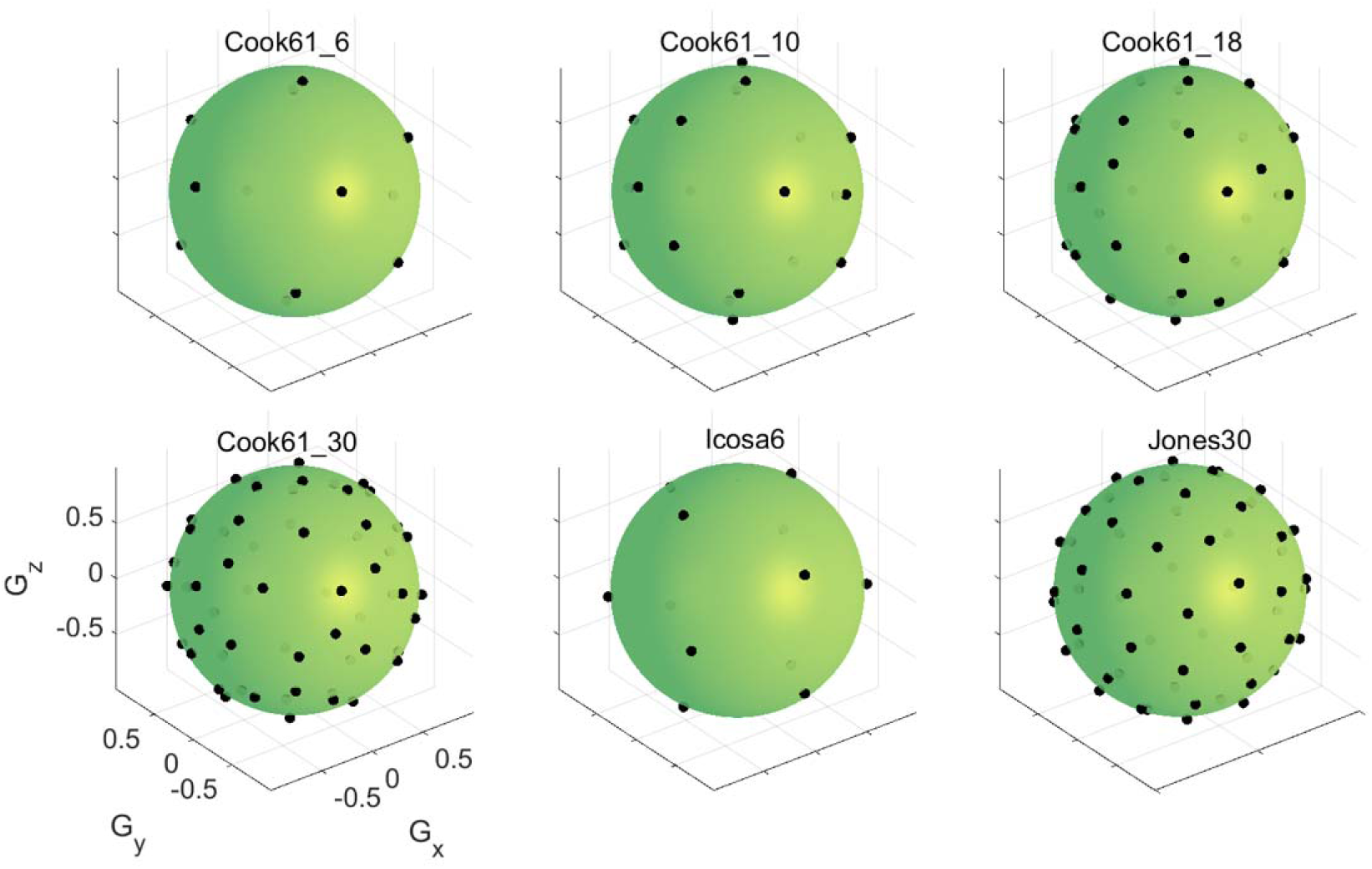
Diffusion encoding schemes investigated include Cook61 diffusion encoding scheme subsampled to 6, 10, 18 and 30 directions, a 6-direction icosahedral scheme and a 30-direction scheme by Jones et al. Each direction was reflected on the opposite side of the sphere for illustration purposes.

The full set of data acquired included NR(ND_orth3, b50_) = 24, NR(ND_Cook61_30, b500_) = 12, NR(ND_Cook61_6, b500_) = 8, NR(ND_icosa6, b500_) = 20 and NR(ND_Jones30, b500_) = 4. This yielded a total of number of acquisitions (NA_all_) = 24 × 3 + 12 × 30 + 8 × 6 + 20 × 6 + 4 × 30 = 720 volumes, which constituted the reference dataset. The nominal acquisition time = NA_all_ × TR = 36 min @ 60 bpm. The reference dataset was subsampled into the 6 diffusion encoding schemes listed in Figure 1, with the total number of high b-value acquisitions (NA_b500_) set to 30, 60, 90 and 120 volumes each. For a 6-direction scheme, this translated to NR = 5, 10, 15 and 20 repetitions respectively. To keep acquisition times consistent, schemes with larger ND had proportionally lower NR. In cases where NA was not divisible by ND e.g. Cook61_18, the last repetition of data would form an incomplete shell. Further to the diffusion encoding schemes, we also investigated the sensitivity of DTI to low b-value acquisitions. Here, we sampled a number of low b-value acquisitions (NA_b50_) equal to a factor of the number of high b-value acquisition (NA_b500_), where NA_b50_ = NA_b500_ / [10, 5, 3, 2]. For example, in a 6- direction scheme with NA_b500_ = 30, NA_b50_ = 3, 6, 10 and 15. The total number of acquisitions (NA_all_) was the sum of NA_b50_ and NA_b500_. This yielded 96 subsampled combinations i.e. acquisition schemes comprising 6 diffusion encoding schemes, 4 sets of low b-value acquisitions and 4 sets of high b-value acquisitions, as reflected in Table 1. The naming convention follows the format “DES_NA_b500__NA_b50_”, e.g. Cook61_30_90_30. Nominal acquisition time was equal to NA_all_ multiplied by number of slices, assuming a heart rate of 60 beats per minute (bpm).

**Table 1.** Acquisition schemes extracted from the reference dataset, indicating numbers of diffusion encoding directions for high b-value data (ND_b500_), numbers of acquisition volumes for high b-value data (NA_b500_), numbers of repetitions for high b-value data (NR_b500_), numbers of acquisition volumes for low b-value data (NA_b50_) including three orthogonal diffusion encoding directions, total numbers of acquisition volumes (NA_all_), and acquisition time. For brevity and clarity, subsets of data with different diffusion encoding schemes (highlighted yellow) and NA_b50_ (highlighted blue) are reported in detail in Results.

| Diffusion encoding scheme |  | Cook61_6 / Icosa6 |  |  |  | Cook61_10 |  |  |  | Cook61_18 |  |  |  | Cook61_30 / Jones30 |  |  |  |
| --- | --- | --- | --- | --- | --- | --- | --- | --- | --- | --- | --- | --- | --- | --- | --- | --- | --- |
| $ND_{b500}$ | | 6 | 6 | 6 | 6 | 10 | 10 | 10 | 10 | 18 | 18 | 18 | 18 | 30 | 30 | 30 | 30 |
| $NA_{b500}$ | | 30 | 60 | 90 | 120 | 30 | 60 | 90 | 120 | 30 | 60 | 90 | 120 | 30 | 60 | 90 | 120 |
| $NR_{b500}$ | $= NA_{b500} / ND_{b500}$ | 5 | 10 | 15 | 20 | 3 | 6 | 9 | 12 | 1.7 | 3.3 | 5 | 6.7 | 1 | 2 | 3 | 4 |
| $NA_{b50}$ | $= NA_{b500} / [10, 5, 3, 2]$ | 3 | 6 | 9 | 12 | 3 | 6 | 9 | 12 | 3 | 6 | 9 | 12 | 3 | 6 | 9 | 12 |
|  |  | 6 | 12 | 18 | 24 | 6 | 12 | 18 | 24 | 6 | 12 | 18 | 24 | 6 | 12 | 18 | 24 |
|  |  | 10 | 20 | 30 | 40 | 10 | 20 | 30 | 40 | 10 | 20 | 30 | 40 | 10 | 20 | 30 | 40 |
|  |  | 15 | 30 | 45 | 60 | 15 | 30 | 45 | 60 | 15 | 30 | 45 | 60 | 15 | 30 | 45 | 60 |
| $NA_{all}$ | $= NA_{b50} + NA_{b500}$ | 33 | 66 | 99 | 132 | 33 | 66 | 99 | 132 | 33 | 66 | 99 | 132 | 33 | 66 | 99 | 132 |
|  |  | 36 | 72 | 108 | 144 | 36 | 72 | 108 | 144 | 36 | 72 | 108 | 144 | 36 | 72 | 108 | 144 |
|  |  | 40 | 80 | 120 | 160 | 40 | 80 | 120 | 160 | 40 | 80 | 120 | 160 | 40 | 80 | 120 | 160 |
|  |  | 45 | 90 | 135 | 180 | 45 | 90 | 135 | 180 | 45 | 90 | 135 | 180 | 45 | 90 | 135 | 180 |
| Time (s) | $= N_{all} * 3R-R$<br>intervals<br>(= 3s @ 60bpm) | 99 | 198 | 297 | 396 | 99 | 198 | 297 | 396 | 99 | 198 | 297 | 396 | 99 | 198 | 297 | 396 |
|  |  | 108 | 216 | 324 | 432 | 108 | 216 | 324 | 432 | 108 | 216 | 324 | 432 | 108 | 216 | 324 | 432 |
|  |  | 120 | 240 | 360 | 480 | 120 | 240 | 360 | 480 | 120 | 240 | 360 | 480 | 120 | 240 | 360 | 480 |
|  |  | 135 | 270 | 405 | 540 | 135 | 270 | 405 | 540 | 135 | 270 | 405 | 540 | 135 | 270 | 405 | 540 |

### Data Analysis

Post-processing used in-house tools developed in Python. Image registration was performed by masking a suitable low b-value image using a square, registering all b_low_ images to this reference image, then using the average of registered b_low_ images as a reference image to register all images. The DTI signal representation was then fitted to the full dataset using robust weighted least squares^39^ implemented into DiPy^40^, and the entire image series was predicted from this fit. All original images were then re-registered to these predicted images, leading to superior registration compared to the first stage. Registration was performed using SimpleITK^41^ with Mutual Information as a metric, calculated within the square mask. Diffusion tensor fitting was performed on the registered images using robust weighted-least squares^39^. The robust fitting method has been previously shown to be superior to whole-image shot-rejection^39^. cDTI parameters MD, FA, helix angle (HA) and sheetlet angle (E2A) were calculated. HA and E2A were measured using a cylindrical coordinate system with origin at the centre of mass of the left ventricular (LV) segmentation on a slice-wise basis, as defined here^42^. Segmentation of the LV contours was performed with care taken to exclude voxels exhibiting partial-volume effects. Regions affected by strong artefacts that may negatively impact the results, were masked out by defining ‘sectors’ centred on the LV blood-pool such that these voxels are ignored in the voxel statistics.

Bootstrapping was done using the repetition bootknife method^43^ which is a form of stratified bootstrapping. Each diffusion encoding direction was treated as a strata, and each bootstrap sample was generated by randomly choosing (with replacement) images from each strata, after first removing a random image from each strata. A total of 500 bootstrap samples were generated per acquisition scheme (Table 1). This was well in excess of the minimum number of bootstrap samples required for stable measurements of accuracy and precision, and in a similar range as in the previous literature^44,45^. The number of images chosen from each strata was based on the number of repetitions specified for each shell. Where NA_b500_ was perfectly divisible by ND_b500_, i.e. in most cases, these images were distributed equally across diffusion encoding directions for the current shell. Where NA_b500_ was not perfectly divisible by ND_b500_ (e.g. in the Cook61_18 scheme with NA_b500_ = 30), any remaining images were then assigned to random unique directions within the design for the current shell, so there was at most a difference of 1 image per strata in the current shell.

For each individual diffusion encoding scheme, accuracy was assessed by calculating the root mean squared difference (RMSD) between the bootstrap samples and the full reference dataset. Precision was assessed by the standard deviation (SD) of cDTI metrics across bootstrap samples, whereby a lower SD reflected higher precision, i.e. precision = (SD)^-2^. The mean, RMSD and SD were then averaged over voxels in segmented regions-of-interest (ROIs) in a mid-myocardial slice. Boxplots of mean, RMSD and SD are presented with median and interquartile range (IQR) over subjects. Individual bootstrap sample data in all volunteers from selected diffusion encoding schemes were also presented as histograms. The data were fitted using normal distributions. Non-overlapping 95% confidence intervals (CI) of the mean indicate statistically significant differences between acquisition schemes. Paired t-tests for RMSD and SD measures were performed between each acquisition scheme. P-values were adjusted for multiple comparisons using Bonferroni-Holm correction^46^ with a significance threshold of p < 0.05. P-value matrices were calculated, and cDTI parameters compared against nominal imaging time.

## RESULTS

MD, FA, HA and E2A maps in a representative volunteer are shown (Figure 2). The maps are consistent with those reported in the literature. Absolute differences with respect to the reference data and SD were elevated in the inferolateral wall, corresponding to the region near the posterior vein, but this effect was less distinct in the average parameter maps.

**Figure 2.**
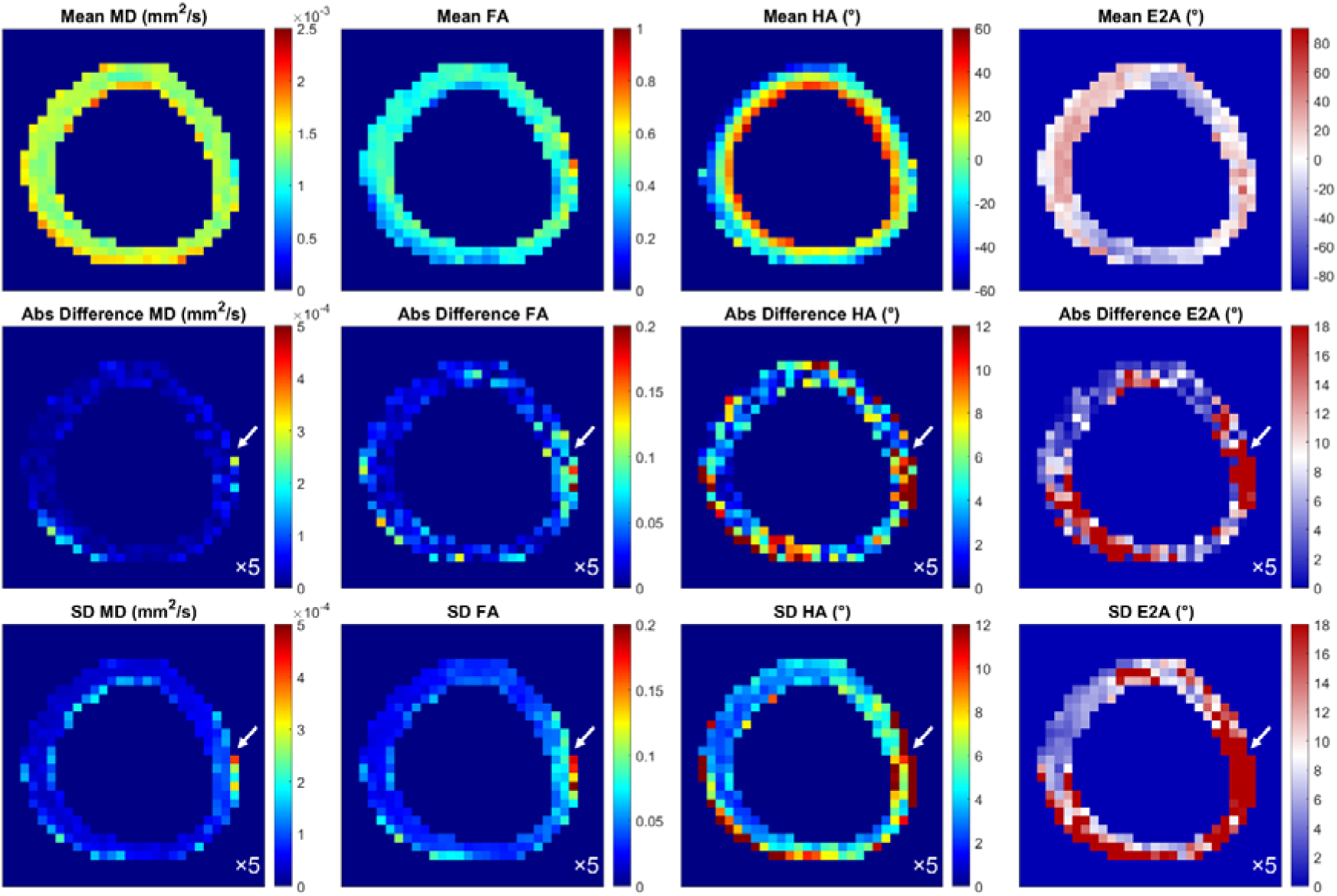
cDTI maps MD, FA, HA and E2A in a mid-myocardial short-axis slice in a representative healthy volunteer using example diffusion encoding scheme Cook61_30 with NA_b500_ = 90 and NA_b500_ = 30. (Top) mean over bootstrap samples, (middle) absolute difference between current and reference diffusion encoding scheme and (bottom) standard deviation across bootstrap samples. Region of elevated absolute difference and SD in the inferolateral wall is indicated by arrows. These maps were scaled at 5× smaller range to highlight the heterogeneity.

Boxplots reflecting (i) cDTI metrics averaged over bootstrap samples, (ii) accuracy of cDTI metrics expressed as the RMSD with respect to the fully sampled reference data, and (iii) 1 / sqrt(precision) as expressed by SD across bootstrap samples across a mid-myocardial short-axis slice are shown (Figure 3; median and IQR over subjects). For clarity of presentation, we focused first on 24 acquisition schemes with 6 different diffusion encoding schemes and 4 different total acquisition times. In all cases, NA_b500_:NA_b50_ = 3. For context, these schemes are highlighted in yellow in Table 1. In the reference data, mean MD = (1.46 ± 0.04) × 10^-3^ mm^2^/s, mean FA = 0.35 ± 0.02, mean HA = -3.4° ± 3.0°, mean E2A = 1.7° ± 7.1° (mean ± SD across subjects). Within each band with normalised acquisition times, there was a small but observable trend towards lower RMSD and SD in MD, FA, HA and E2A with higher ND, indicating higher accuracy and precision. This was consistent across Cook, Icosa and Jones diffusion encoding schemes. For the same acquisition time, for the ND = 30 schemes (Cook and Jones), RMSD MD, FA, HA and E2A were on average 48%, 40%, 34% and 34% lower than for the ND = 6 schemes (Cook and icosahedral). Similarly, SD MD, FA, HA and E2A were 6.2%, 7.4%, 10% and 5.6% lower in the former compared to the latter. Mean FA was most sensitive to diffusion encoding scheme, whilst MD, HA and E2A were less so.

**Figure 3.**
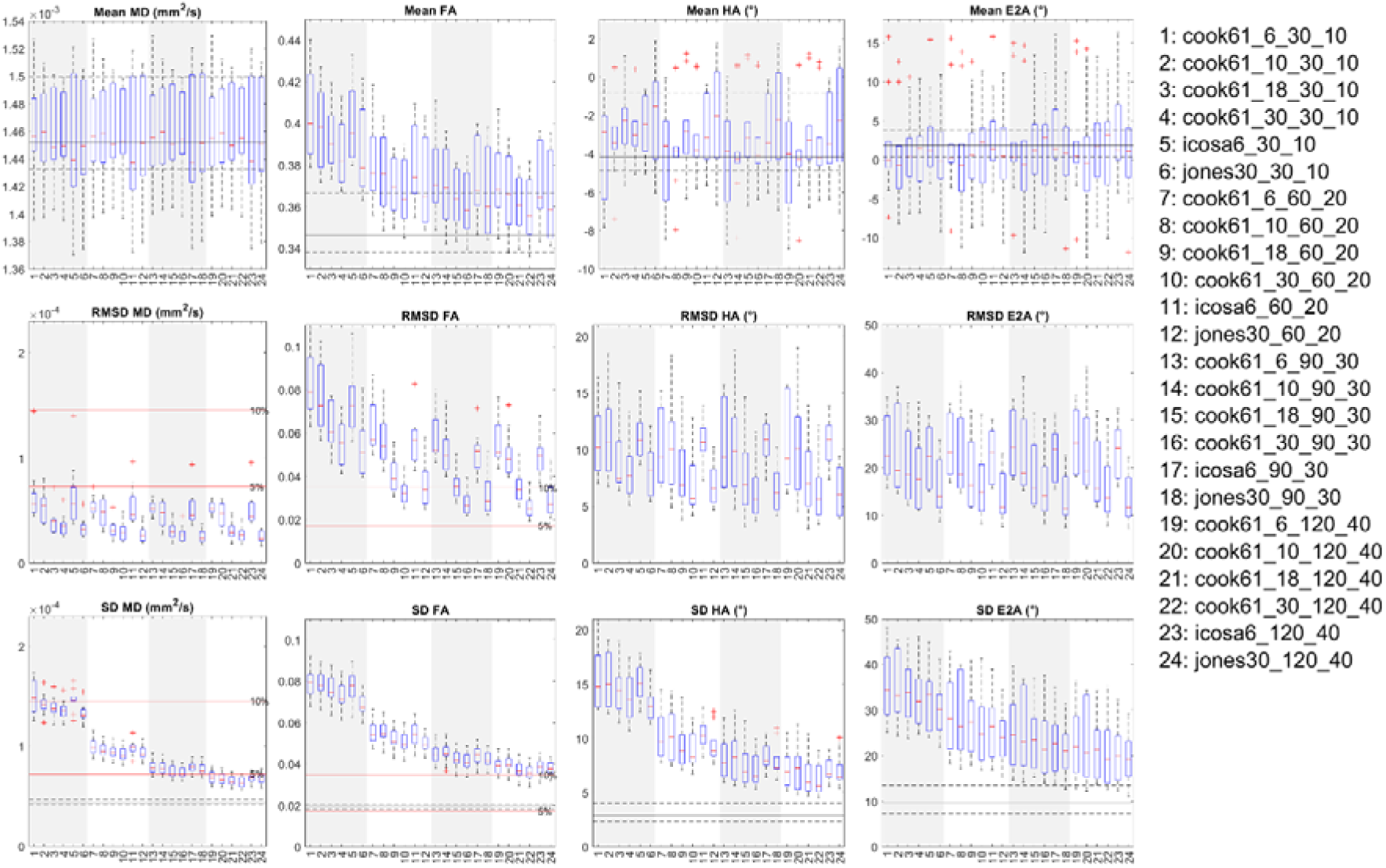
Boxplots of cDTI metrics (left to right) MD, FA, HA and E2A showing (top) cDTI metrics averaged over bootstrap samples, (ii) RMSD with respect to the fully sampled reference data, and (iii) SD across bootstrap samples across a mid-myocardial short-axis slice. 24 acquisition schemes are described in the following format “DES_NA_b500__NA_b50_”, e.g. Cook61_30_90_30. These were sorted by diffusion encoding scheme and grouped into four groups (white and grey vertical bands) with increasing NA_all_ corresponding to increasing acquisition times. For reference, median and IQR values from the reference dataset are given (black solid and dashed lines); 5% and 10% of the median MD and FA from the reference dataset are indicated (red solid lines).

Across bands with different acquisition times, there was a clear trend towards lower SD (i.e. better precision) in all cDTI metrics with increasing number of acquisitions and scan time. For the NA_all_ = 160 schemes, SD MD, FA, HA and E2A were on average 53%, 50%, 52% and 36% lower than for the NA_all_ = 40 schemes. Similarly, RMSD in FA (but not other parameters) was lower with increasing number of acquisitions and scan time, indicating better accuracy. For ease of reference, these results were reformatted into bands with different diffusion encoding schemes and ordered by increasing NA_all_ within each band (Supplementary Figures 1, 2 and 3).

A subset of six acquisition schemes corresponding to NA_all_ = NA_b500_ + NA_b50_ = 120 are presented as histograms (Figure 4). RMSD and SD for MD, FA, HA and E2A were significantly lower for ND ≥ 18 compared to ND = 6 data, indicating better accuracy and precision in diffusion encoding schemes with greater number of directions rather than repetitions. This applied across Cook, icosahedral and Jones diffusion schemes. P-value matrices illustrate significant differences (p < 0.05) between acquisition schemes, that were most prominent in precision (SD) and least prominent in mean values across different ND and NA (Figure 5).

**Figure 4.**
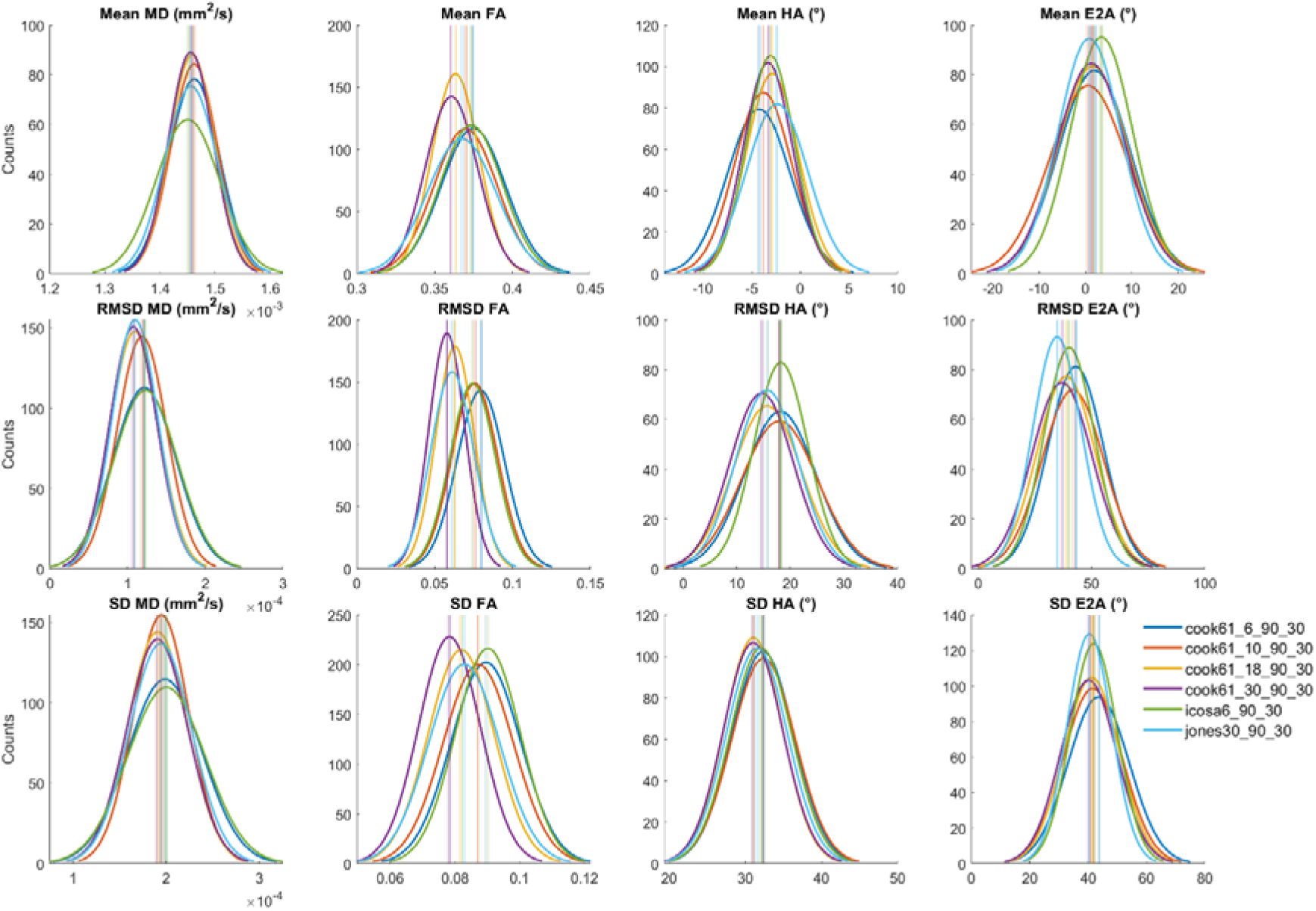
Histograms of (top to bottom) mean, RMSD and SD of (left to right) MD, FA, HA and E2A across 500 bootstrap samples and healthy volunteers (N = 10). Data from six time-normalised acquisition schemes are presented, with vertical lines indicating 95% confidence intervals of the mean. Non-overlapping 95% CI indicate significant differences between groups.

**Figure 5.**
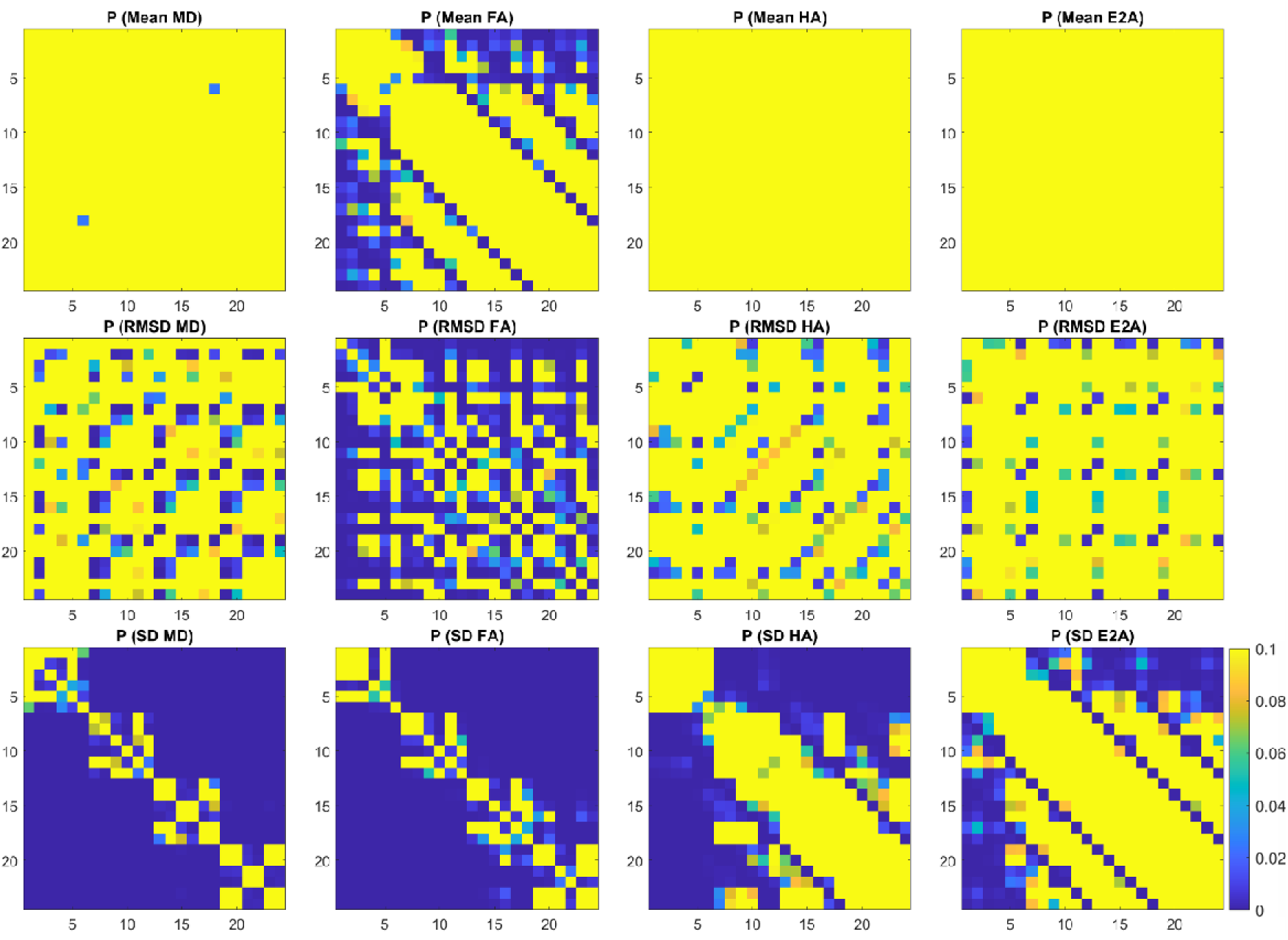
P-value matrices reflecting pairwise comparisons between the 24 acquisition schemes given in Figure 3, with p < 0.05 indicating significant differences. Intensity range was scaled between 0 and 0.1 to highlight results close to p = 0.05.

We also report the mean, RMSD and SD for the four cDTI parameters as a function of numbers of low and high b-value acquisitions, NA_b50_ and NA_b500_. For clarity, only a single diffusion encoding scheme Cook61_30 is presented (Figure 6). For context, these schemes are highlighted in blue in Table 1. Within each band of fixed NA_b500_, there was a trend towards lower RMSD and SD of MD and FA. A similar trend was observed in RMSD and SD of HA and E2A at lower total number of acquisitions (NA_all_ < 45), but was not discernible at higher NA_all_. For the same NA_b500_, the schemes with NA_b500_:NA_b50_ = 2 had on average, RMSD MD and FA that were 28% and 11% lower than the NA_b500_:NA_b50_ = 10 schemes. Similarly, SD MD and FA were 46% and 19% lower in the former compared to the latter.

**Figure 6.**
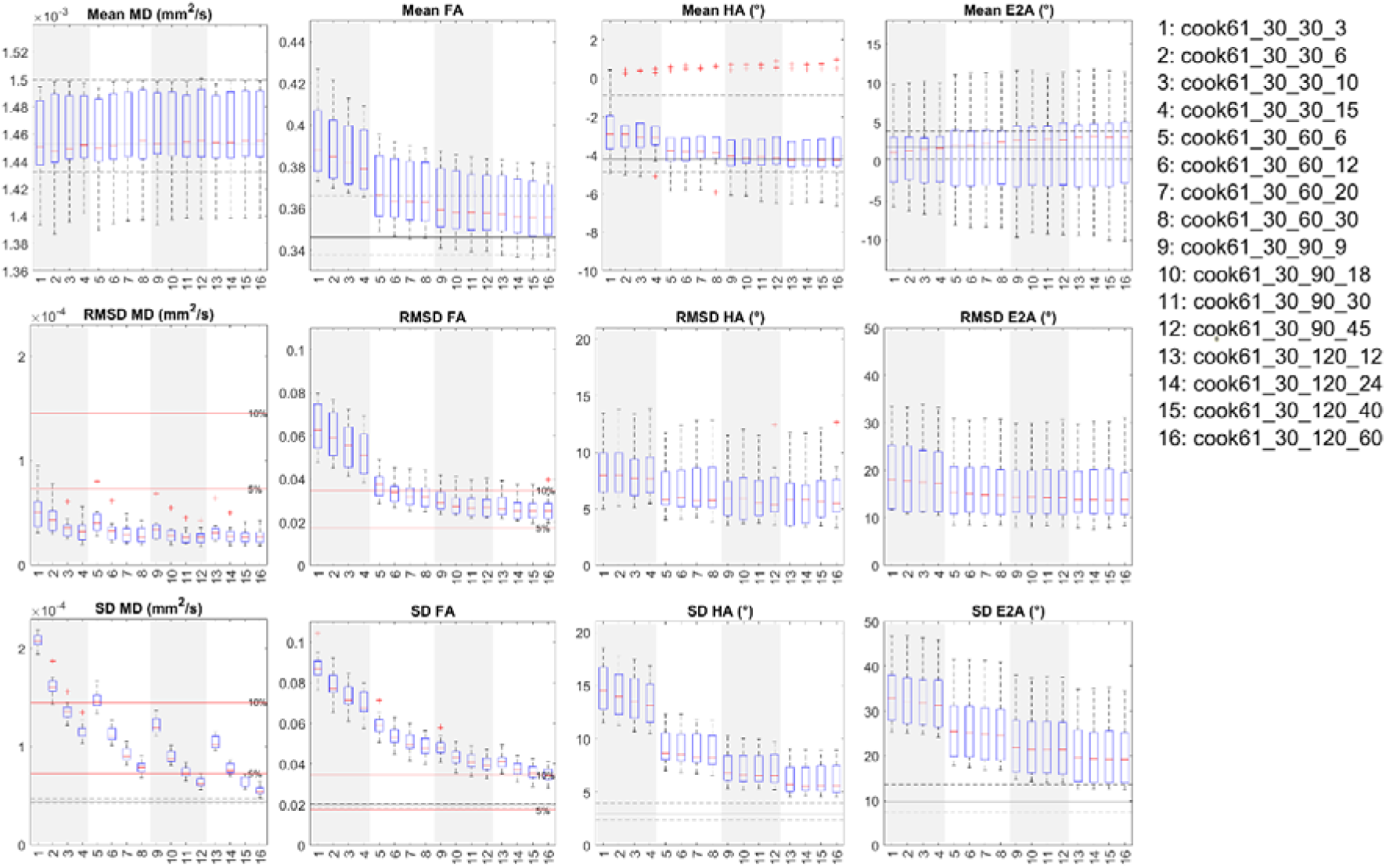
Boxplots of cDTI metrics (left to right) MD, FA, HA and E2A showing (top) cDTI metrics averaged over bootstrap samples, (ii) RMSD with respect to the fully sampled reference data, and (iii) SD across bootstrap samples across a mid-myocardial short-axis slice. 16 acquisition schemes were sorted by number of low b-value acquisitions (NA_b50_) and grouped into four groups (white and grey vertical bands) with increasing NA_all_ corresponding to increasing acquisition times. For reference, median and IQR values from the reference dataset are given (black solid and dashed lines); 5% and 10% of the median MD and FA from the reference dataset are indicated (red solid lines).

A subset of a single diffusion encoding scheme with four sets of NA_b50_ are presented as histograms (Figure 7). RMSD and SD of MD and FA were significantly lower for NA_b50_ ≥ 18 compared to NA_b50_ ≤ 10 data, indicating better accuracy and precision with greater number of low b-value acquisitions. No significant differences were observed in RMSD and SD of HA and E2A. P-value matrices illustrate significant differences (p < 0.05) that were most prominent in accuracy (RMSD) and precision (SD) of MD and FA across different NA_b50_ and NA_all_ (Figure 8).

**Figure 7.**
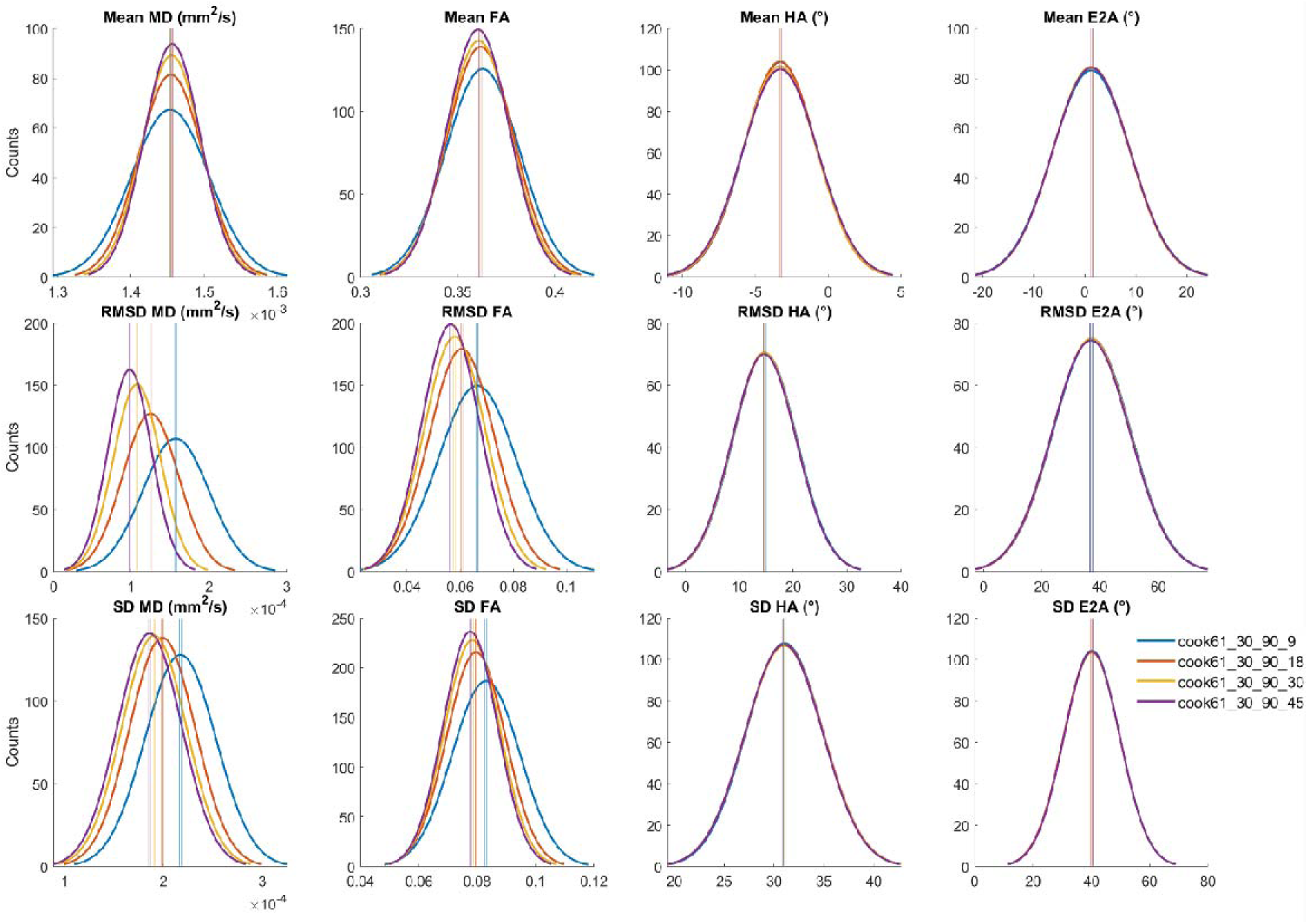
Histograms of (top to bottom) mean, RMSD and SD of (left to right) MD, FA, HA and E2A across 500 bootstrap samples and healthy volunteers (N = 10). Data from a single diffusion encoding scheme with different numbers of low b-value acquisitions (NA_b50_ = 9, 18, 30, 45) are presented, with vertical lines indicating 95% confidence intervals of the mean. Non-overlapping 95% CI indicate significant differences between groups.

**Figure 8.**
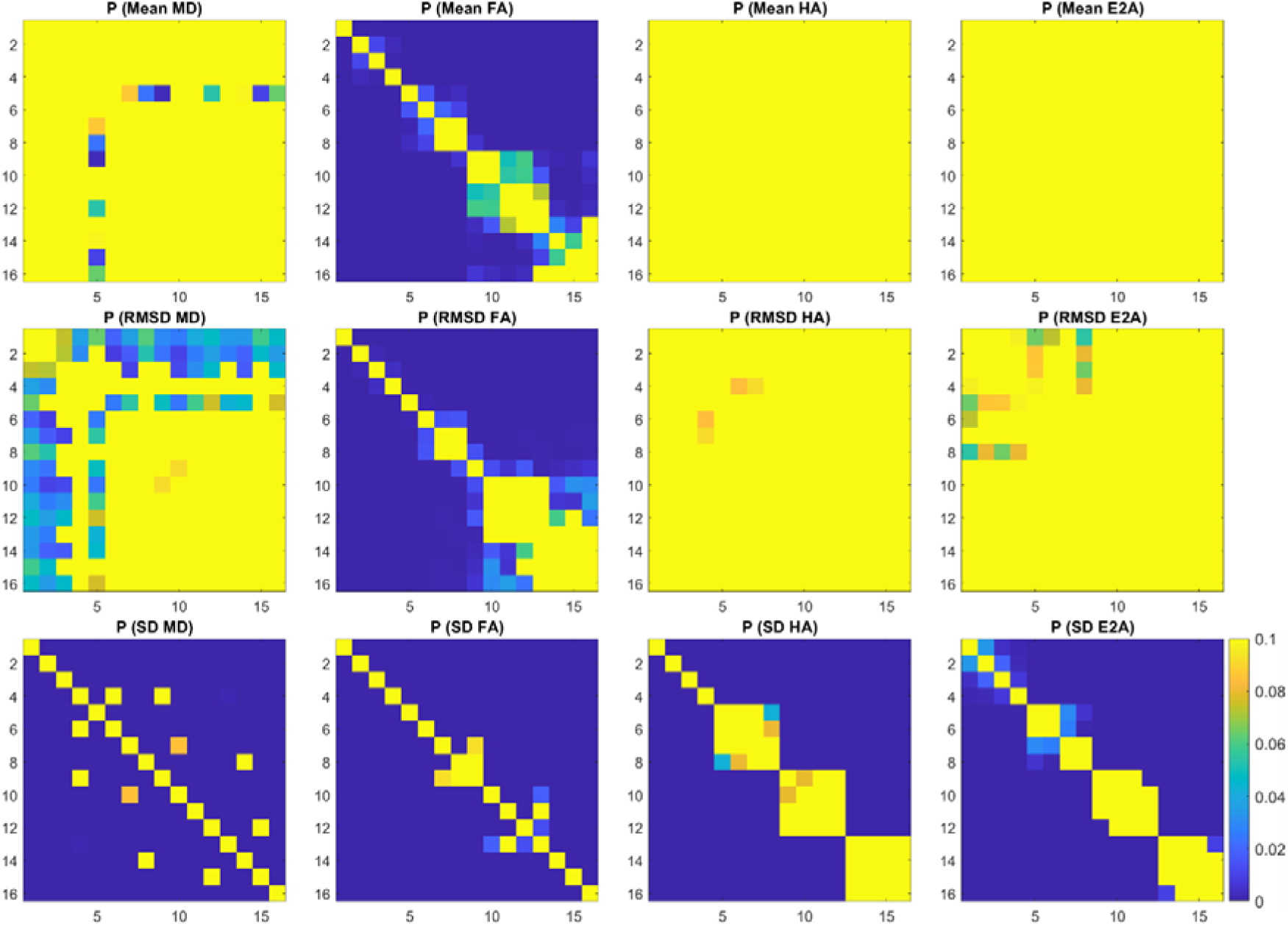
P-value matrices reflecting pairwise comparisons between the 16 acquisition schemes given in Figure 6, with p < 0.05 indicating significant differences.

Average mean, RMSD and SD of cDTI parameters were evaluated as a function of acquisition time (Figure 9). The data show decreasing RMSD and SD with increasing NA_all_ and scan time. The majority of diffusion encoding schemes investigated yielded an RMSD MD of <5% and SD MD of < 10% of the reference MD. Acquisition schemes DES_All__90_45, DES_All__120_40 and DES_All__120_60 yielded SD MD of < 5% of the reference MD, where DES_All_ corresponded to all diffusion encoding schemes. Differences in MD between time-normalised diffusion encoding schemes were marginal. Diffusion encoding schemes were ranked from most to least accurate FA (i.e. lowest to highest RMSD): Cook61_30, Jones30, Cook61_18, Icosa6, Cook61_10, Cook61_6, with RMSD FA < 10% of the reference FA for Cook61_30 / Jones30 with NA_all_ ≥ 108. SD FA < 10% of the reference FA for Cook61_30_120_60 only. Diffusion encoding schemes were ranked (i) from most to least accurate HA: Cook61_30, Jones30, Cook61_18, Cook61_10, Cook61_6, Icosa6, (ii) from most to least precise HA: Cook61_30, Cook61_18, Jones30, Icosa6, Cook61_10, Cook61_6, (iii) from most to least accurate E2A: Jones30, Cook61_30, Cook61_18, Icosa6, Cook61_10, Cook61_6, and iv) from most to least precise E2A: Jones30, Icosa6, Cook61_30, Cook61_18, Cook61_6, Cook61_10. NA_b500_ = 120 and Cook61_30 / Cook61_18 was needed to achieve SD HA < 7°, whilst SD E2A < 21° was achievable with NA_b500_ = 120 and Jones30, Icosa6.

**Figure 9.**
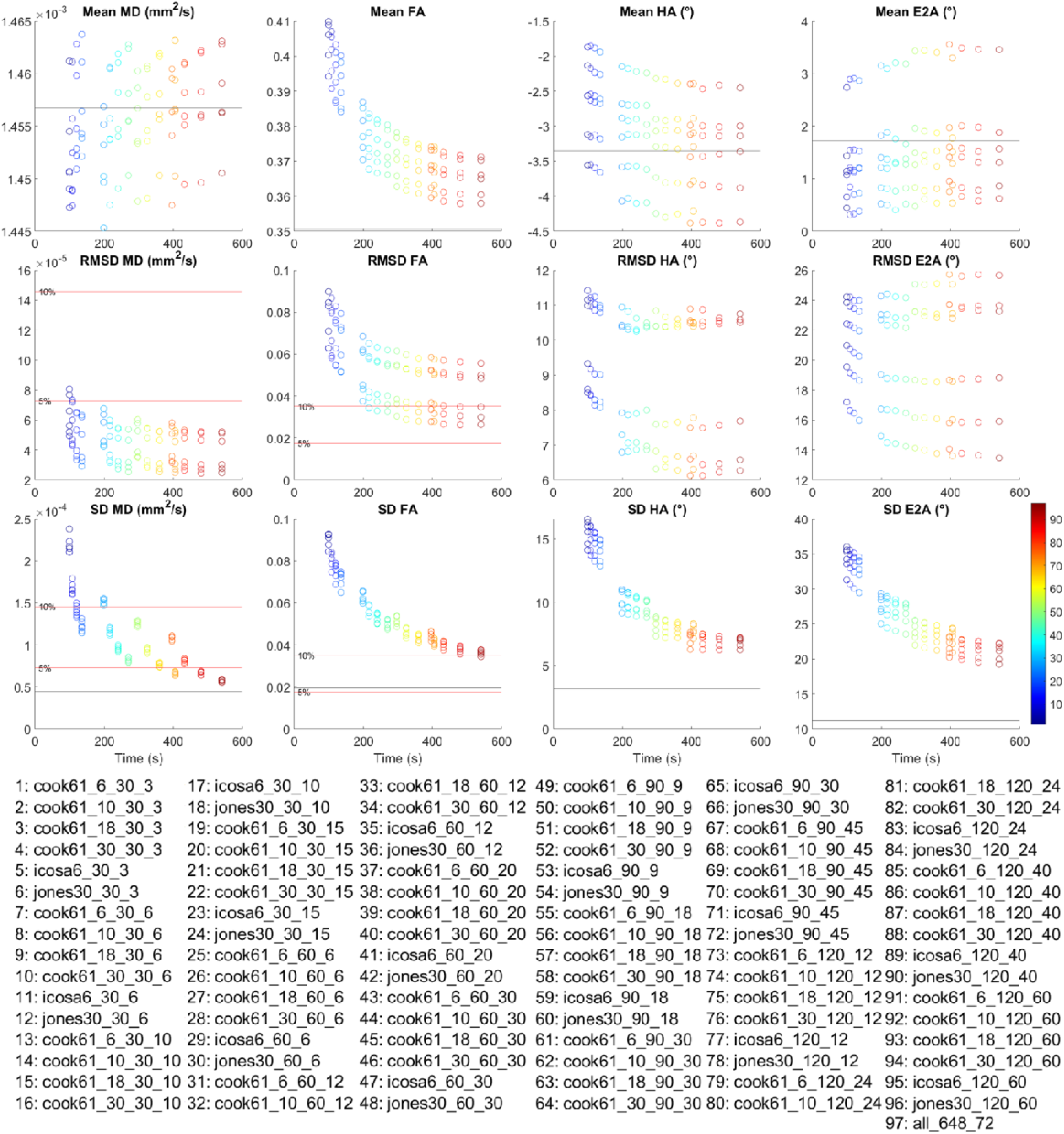
Mean, RMSD and SD of MD, FA, HA and E2A within a mid-myocardial slice averaged across subjects and plotted against nominal acquisition time. Each acquisition scheme 1 – 97 is encoded by colour and described in the legend. For better presentation, mean values for the reference dataset labelled “all_648_72” are not plotted due to long scan time, and are instead given by black lines; 5% and 10% of the mean MD and FA from the reference dataset are indicated by red lines.

Higher NA_all_ i.e. longer acquisition times led to improved precision in all DTI metrics whereby quadrupling NA_all_ from 40 to 160 volumes led to a 53%, 50%, 52% and 36% reduction in SD in MD, FA, HA and E2A, averaged across six diffusion encoding schemes. In relative terms, this corresponded to a reduction in SD MD from 9.7% to 4.6% and SD FA from 22% to 11%, expressed as a percentage of the mean MD and FA of the reference data. Similarly, accuracy improved with a 21%, 40%, 13% and 4.5% reduction in RMSD in MD, FA, HA and E2A from 40 to 160 acquired volumes. In relative terms, this corresponded to a reduction in RMSD MD from 3.3% to 2.7% and RMSD FA from 19% to 12% of the reference mean MD and FA. Absolute values of RMSD and SD in selected acquisition schemes are given in Table 2.

**Table 2.**
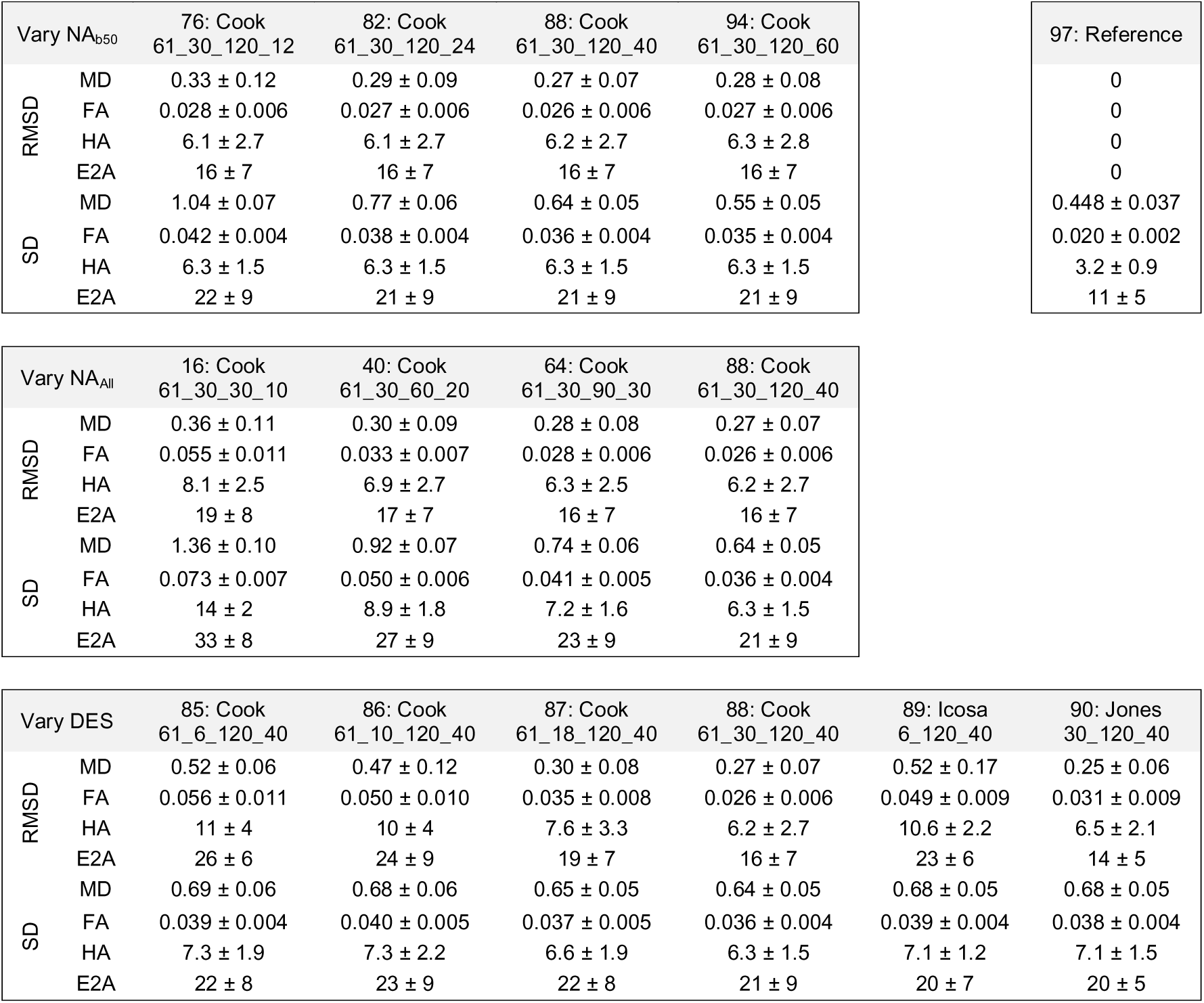
RMSD and SD of MD (×10^-4^ mm^2^/s), FA, HA (°), E2A (°) for selected acquisition schemes, varying (top to bottom) NA_b50_, NA_All_ and diffusion encoding scheme (DES; mean ± SD over subjects). For ease of reference, the acquisition schemes were numbered according to the legend in Figure 9.

## DISCUSSION

We evaluated 96 acquisition schemes in terms of accuracy and precision with respect to the 720-volume i.e. ∼36-minute reference datasets. First, we observed that accuracy and precision in all estimates improved with NA_all_ and acquisition time, consistent with previous reports^30,33^. Second, we found that prioritising ND over NR given a fixed acquisition time improved accuracy and precision in general. Differences in accuracy and precision between different diffusion encoding schemes with equal ND and NA_All_ were less prominent and generally not significant.

In our pilot work in healthy volunteers^33^, we reported RMSD MD = 2.29 (×10^-4^ mm^2^/s), RMSD FA = 0.08 and RMSD HA = 13° using a Jones scheme with ND = 6, NA_b50_ = 24, and NA_b500_ = 96. The accuracy improved to RMSD MD = 1.11 (×10^-4^ mm^2^/s), RMSD FA = 0.04 and RMSD HA = 7.8° when using a Caruyer scheme^47^ with a larger number of directions, i.e. ND = 96 and NR = 1. In the current study, the nearest equivalent acquisition scheme to the former was Cook61_6_90_30 which yielded RMSD MD = 0.794 ×10^-4^ mm^2^/s, RMSD FA = 0.04 and RMSD HA = 8.4°. This represented an improvement in accuracy compared to the previous study, that may be attributed to the improved post-processing pipeline with robust fitting of tensors^39^ over shot rejection. Direct comparison to a 96-direction diffusion encoding scheme was not available. Another pilot study in healthy volunteers (N = 5) explored time-normalised cDTI acquisitions with six diffusion encoding schemes where ND = 6, 10, 12, 15, 20, 30 and NR = 10, 6, 5, 4, 3, 2 respectively^32^. The authors reported minimal differences in median and interquartile intervals of MD and FA, and they suggested that the acquisition scheme was not critical to measuring MD and FA. In a separate study using stimulated echo acquisition mode (STEAM), no significant differences were found in MD and FA between time-normalised averaged datasets where ND = 6, 10, 12 and 20^48^. Our results suggest otherwise and indicate that prioritising ND over NR for a given scan time, up to ND = 30, improves accuracy and precision of cDTI parameters. For example, RMSD and SD in MD and FA were significantly lower in Cook61_30_120_40 compared to Cook61_6_120_40 (p < 0.001). Moreover, there is a potential for occasional poor image quality when diffusion encoding is applied in a particular direction e.g. due to eddy current effects. Acquiring the minimum of ND = 6 provides no redundancy in case a specific diffusion encoding direction gives rise to sub-optimal image quality.

The effects of ND have also been reported ex vivo^30,49^. One study in fixed pig hearts found that robust estimation of HA could be obtained with ND/NR = 12/6, 30/3 or 64/2, recommending the 12/6 combination as one with the shortest (<10 min) acquisition time^49^.

However, only qualitative assessments were reported and data were not time-normalised. A second study in fixed rat hearts investigated the effects of ND, SNR and spatial resolution on the accuracy and precision of cDTI^30^. It was shown that for a given scan time, the precision of FA, HA, E2A, transverse angle, sheetlet elevation and sheetlet azimuth for a given scan time were largely independent of the choice of increasing NR or ND, on the assumption that NR is proportional to √SNR. In practice, physiological effects in vivo, such as residual motion effects arising from breathing and cardiac contraction, contribute a significant additional noise-like component that violates the above assumption, resulting in spatially correlated variations in parameters. This may, to an extent, reduce the value of additional NR, and support our current findings of prioritising ND over NR, up to ND = 30. Extrapolating to typical parameter settings e.g. resolution used in the clinic, the study suggested that the expected bias in MD and FA were 2.1% and 13% and precision in MD and FA were ±4.9% and ±30% with respect to the ground truth; precision of HA and E2A were ±14°and ±24° respectively. Direct comparisons between ex vivo and in vivo data are difficult due to various reasons such as different physiological status and temperature of the myocardium. Nonetheless, our current findings similarly reflect that precision of MD is superior to that of FA, and precision of HA is superior to that of E2A, although the specific values differ. Whilst a ground truth does not exist in vivo, we can see that the trends in accuracy and precision appear to approach an asymptotic value by NA_All_ = 160, justifying the appropriateness of NA_All_ = 720 as suitable reference data.

In the above rat heart study, precision in MD was optimised by maximising NR of non-diffusion-weighted scans at the expense of ND, for a given scan time. This leads us to our third observation that accuracy and precision of MD, and to a lesser extent FA, improved with increasing numbers of low b-value acquisitions NA_b50_, with disproportionately large benefits at low NA_All_. With larger NA_All_ ≥ 99, as more commonly used in the clinic, improvements in both accuracy and precision in MD and FA were retained. For example, with diffusion encoding scheme Cook61_30, NA_b500_ = 90 and NA_b50_ = [9, 18, 30, 45], the improvements in precision of MD and FA between each increasing step of NA_b50_ were significant (p < 0.001). Accuracy in MD improved when NA_b50_ increased from 9 to 18 (p < 0.05), but was not significantly better between successive increments of higher NA_b50_.

Other studies investigated the sensitivity of cDTI to SNR in human hearts^45^ and in fixed rat hearts^50^. In the former, systolic median 95% CI of the 1^st^, 2^nd^ and 3^rd^ eigenvectors (**v_1_**, **v_2_** and **v_3_**) were 15.5°, 31.2°, and 21.8°. The authors concluded that precision improved with increasing SNR, but the improvements were minimal beyond NR = 10 corresponding to a 10 min scan. This was consistent with our findings. The latter study reported that mean 95%CI of **v_1_**, **v_2_** and **v_3_** was 3.7°, 10.9° and 10.6° respectively. The poorer precision in vivo may reflect the additional challenges of motion and lower spatial resolution in vivo.

The recommended protocols for cDTI will depend on several factors including the precision of the measurement, the expected differences e.g. between health and pathology, the parameters of interest, subject compliance and scan time available. Despite our finding that higher NA_All_ improved precision, this improvement comes with diminishing returns at higher NA_All_. Moreover, longer scans e.g. >10 min can be challenging to perform in a clinical setting due to limited scan times and increased likelihood of patient discomfort leading to greater patient motion, poorer image quality and premature termination of scans. For context, we consider the range of cDTI parameters seen in clinical cohorts. Where the differences between health and disease are known and expected to be small, the acquisition would need to be designed with greater precision. In general, studies report higher MD and lower FA in disease cohorts relative to controls. In a study of patients post-myocardial infarction^2^, MD was 14% higher, FA was 31% lower and HA was between -7° to +5° relative to controls. In patients with HCM^7,8,10,11^, MD was between 2-10% higher, FA was between 6-17% lower relative to controls. Patients with amyloidosis had 26% higher MD and 29% lower FA relative to controls^13^. Similarly, patients with aortic stenosis had 6% higher MD and 17% lower FA relative to controls^12^. HA and E2A differences in pathology have been primarily reported in terms of HA slope or absolute E2A^2,7,8,10,12,51^ rather than differences in HA and E2A, and may not be directly comparable. Other studies that reported E2A were based on STEAM^3,9^, which is known to yield substantially different measurements compared to spin echo.

In order to achieve SD MD of < 5% of the reference mean, NA_b50_ ≥ 45 with any diffusion encoding scheme was required. This included DES_All__90_45, DES_All__120_40 and DES_All__120_60 where DES_All_ refers to any of six diffusion encoding schemes evaluated. DES_All__90_45 had the shortest acquisition time of 6:45 min. In contrast, only Cook61_30_120_60 yielded SD FA of < 10% of the reference i.e. 9.8%, corresponding to an acquisition time of 9:00 min. Other sequences e.g. Cook61_30_120_40 came close with SD FA = 10.2% with an acquisition time of 8:00 min. That SD FA > SD MD was consistent with the greater sensitivity of FA to noise. SD HA and SD E2A were more sensitive to NA_All_ and diffusion encoding scheme, and less so to NA_b50_. Besides extending the acquisition time, precision in HA and E2A could be improved by prioritising ND over NR, e.g. ND = 30 gave higher precision than ND = 6, given the same total acquisition time. For clinically relevant protocols, our findings support the use of ND = 30. We would also recommend NA_b50_:NA_b500_ ≥ 1/3 for better precision in MD and FA. If only MD were desired, a shorter protocol of < 7 min would be reasonable. We would recommend protocols of > 8 min in order to obtain good precision for FA, HA and E2A, subject to availability of scan time.

## LIMITATIONS

In this study, we have only considered healthy volunteers with good compliance over the entire ∼36 min scan, excluding setup and planning. It is foreseeable that some patients will have poorer compliance leading to poorer image quality, accuracy and precision, suggesting more acquisition volumes and time would be needed to achieve similar levels of accuracy and precision. However, longer acquisition times are likely to contribute to worse compliance and may be infeasible. In the ideal case, we would examine accuracy and precision in a similar study in patients. In practice, such efforts may need to be combined with techniques for accelerating image acquisition, such as simultaneous multi-slice imaging^52^, compressed sensing^53,54^, and deep learning^55–58^.

Image distortion remains a common issue in cDTI. This stems mostly from the use of single shot EPI readouts and consequent sensitivity to susceptibility-induced distortion. This commonly manifests as artefactual compression or dilation of the myocardium, especially near the posterior vein. In this study, we have seen that this effect also contributes to reduced local accuracy and precision. Recent developments in distortion correction using reversed phase encoding data for correction of susceptibility artefacts promise to improve the geometric fidelity of the images^59^.

To maximise the number and scope of acquisition schemes compared, we relied partially on subsampling of the modified Cook61 diffusion encoding scheme. This meant that (i) the subsampled data were not fully independent of each other, and (ii) the distribution of diffusion encoding directions in subsampled schemes was not fully optimal. To mitigate these, fully optimised Icoas6 and Jones30 schemes were employed as independent controls. Their accuracy and precision were seen to be comparable to the Cook61_6 and Cook61_30 schemes respectively. Despite our approach which enabled systematic comparison of a relatively large number of acquisition schemes, we were limited to ND = 30 due to scan time constraints. Some advanced applications e.g. resolving crossing fibres in the brain require larger ND. However, our findings suggest diminishing returns in cDTI with increasing ND > 30, and there is limited evidence for higher ND, particularly where voxels with discrete crossing cell populations are limited in the myocardium. An alternative would be to employ acquisition schemes that are completely independent, as in previous pilot studies^48,49^. However, this approach extends the imaging time, limits the number of acquisition schemes that can be evaluated, and often means that datasets cannot be time-normalised. Even so, the common approach of combining all data to form a reference dataset means that reference data are not completely independent of the component data.

## CONCLUSIONS

In summary, we investigated the trade-off between the number of diffusion encoding directions and repetitions in M2 spin echo-based cardiac DTI. For a given acquisition time, we found that prioritising number of diffusion encoding directions over number of repetitions generally yields better accuracy and precision in cDTI parameters, particularly MD and FA. We also observed that greater sampling of low b-value data improves accuracy and precision of MD and FA but not HA and E2A. Finally, we characterised the improvements in accuracy and precision associated with increasing total number of acquisitions. These results may serve to guide optimisation of protocols for supporting ongoing efforts in harmonisation and standardisation of cDTI and aid its development towards wider clinical adoption.

## Data Availability

The datasets used and/or analysed during the current study are available from the corresponding author on reasonable request.

## LIST OF ABBREVIATIONS

CI: Confidence interval
DES: Diffusion encoding scheme
DTI: Diffusion tensor imaging
E2A: Sheetlet angle
EPI: Echo planar imaging
FA: Fractional anisotropy
HA: Helix angle
IQR: Interquartile range
M2: 2^nd^ order motion compensated
MD: Mean diffusivity
NA: Number of acquisitions
ND: Number of diffusion encoding directions
NR: Number of repetitions
NSA: Number of signal averages
RMSD: Root mean squared difference
SCMR: Society for Cardiovascular Magnetic Resonance
SNR: Signal-to-noise ratio
TE: Echo time
TR: Repetition time

## DECLARATIONS

The authors declare that they have no known competing financial interests or personal relationships that could have appeared to influence the work reported in this paper.

## ETHICS APPROVAL AND CONSENT TO PARTICIPATE

The study was conducted in accordance with the Declaration of Helsinki, and was approved by the UK National Research Ethics Service (19/YH/0324; 18/YH/0168). All subjects provided written informed consent.

## CONSENT FOR PUBLICATION

Not applicable

## AUTHOR CONTRIBUTIONS

Study design and planning (IT), drafting of manuscript (SC, IT), pulse sequence development (CN), data acquisition (IT, DS, RJF), data analysis (SC, IT), clinical oversight and checking for incidental findings (EDA, AMP, NS), funding (IT, JES, EDA, SP), ethics approvals (IT, EDA, SP), review and approval of final manuscript (All).

## ACKNOWLEDGEMENTS

We thank Siemens Healthcare for the pulse-sequence development environment. We thank Dr. Kathryn Richards for her support with study and ethics management.

