## Supplementary Information for "Optimising Cardiac Diffusion Tensor Imaging In Vivo: More Directions or Averages?"

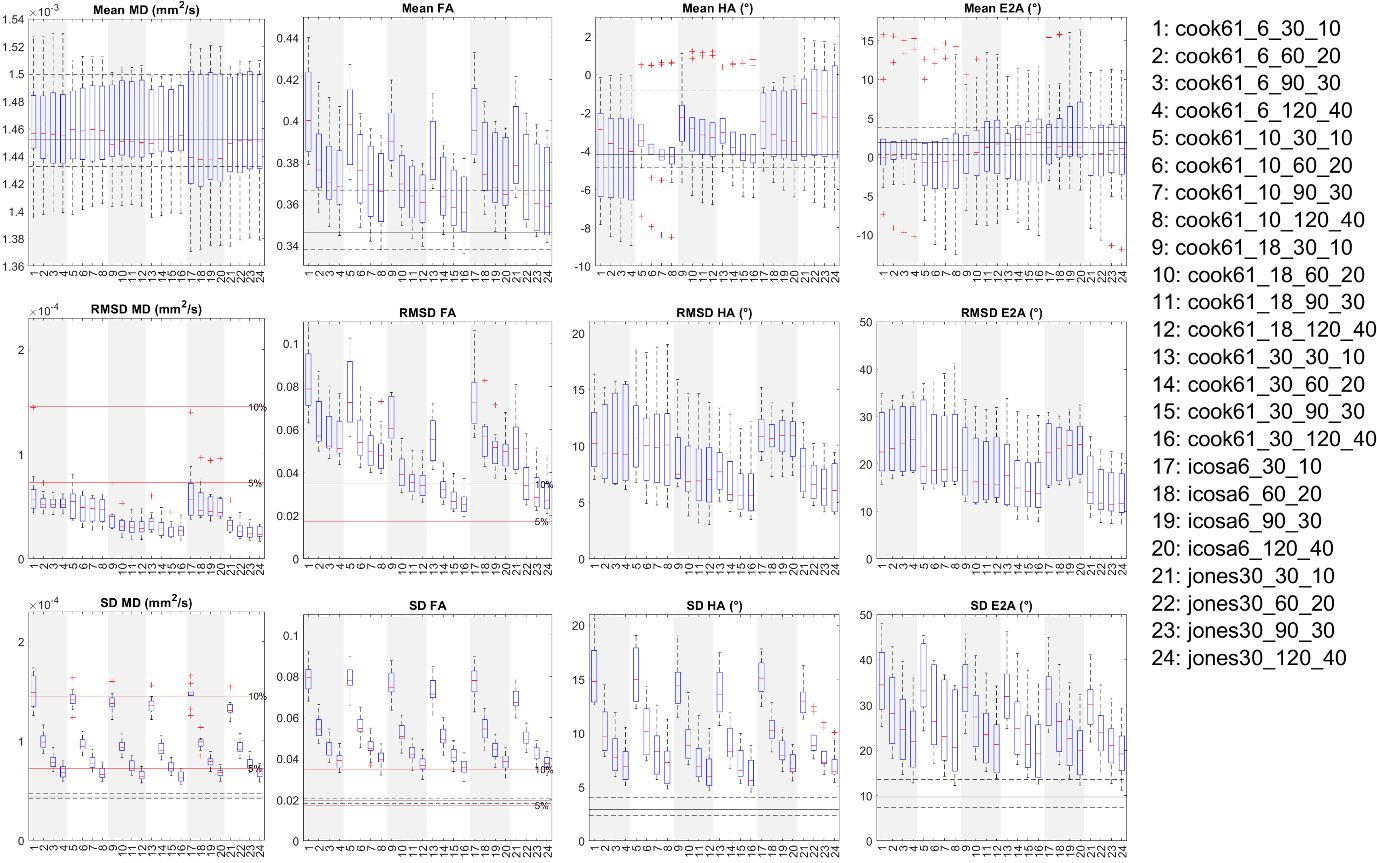
Supplementary Figure 1. Boxplots of cDTI metrics (left to right) MD, FA, HA and E2A showing (top) cDTI metrics averaged over bootstrap samples, (ii) RMSD with respect to the fully sampled reference data, and (iii) SD across bootstrap samples across a mid-myocardial short-axis view. 24 diffusion encoding schemes were sorted by total number of acquisitions (NA_all_) and grouped into six groups (white and grey vertical bands) with different diffusion encoding schemes. For reference, median and IQR values from the reference dataset are given (black solid and dashed lines); 5% and 10% of the median MD and FA from the reference dataset are indicated (red solid lines).

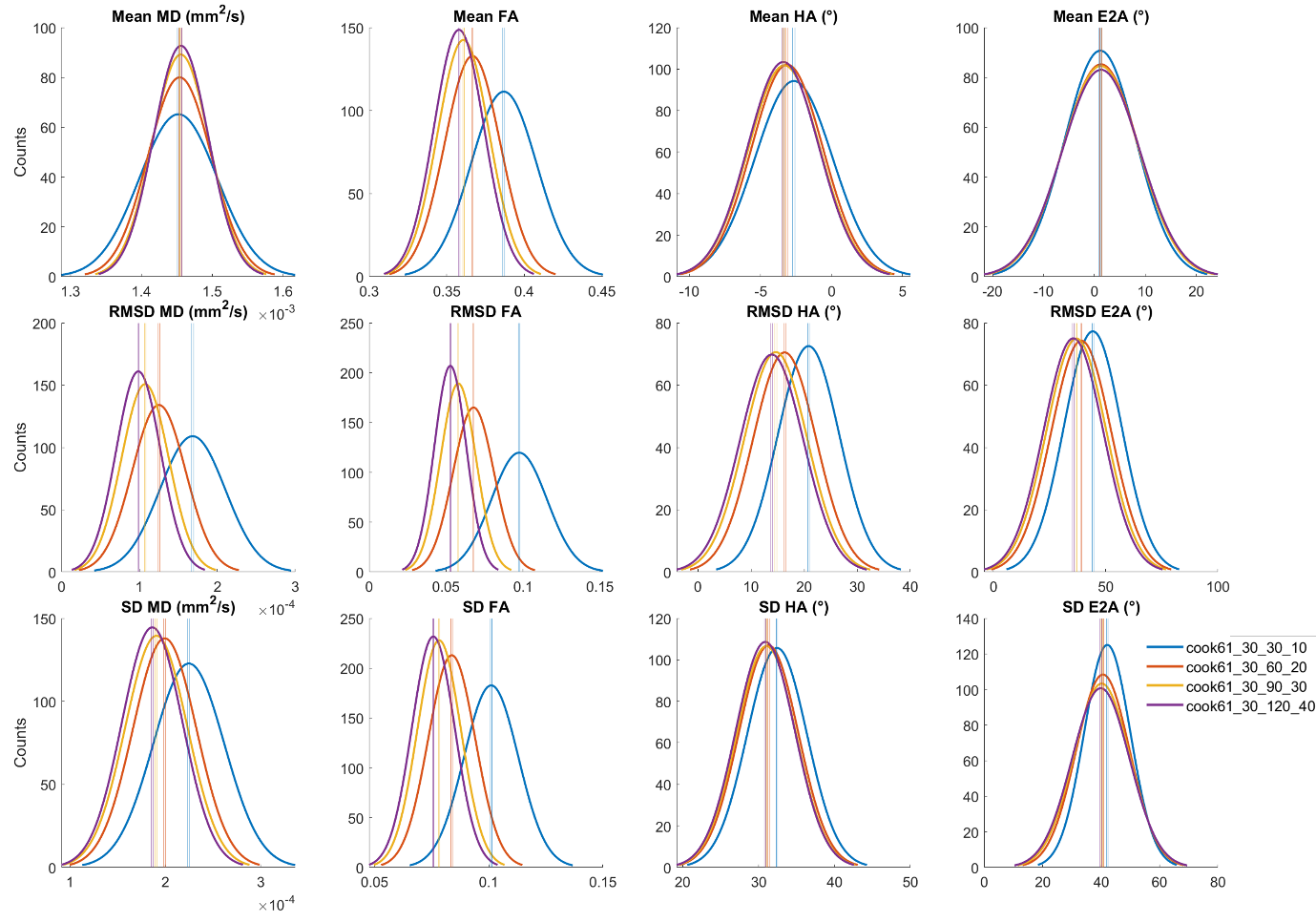

Supplementary Figure 2. Histograms of (top to bottom) mean, RMSD and SD of (left to right) MD, FA, HA and E2A across 500 bootstrap samples and healthy volunteers (N = 10). Data from a single diffusion encoding scheme with different numbers of low and high b-value acquisitions (NA_b500_ = 30, 60, 90, 120; NA_b50_ = 10, 20, 30, 40) are presented, with vertical lines indicating 95% confidence intervals of the mean. Non-overlapping 95% CI indicate significant differences between groups.

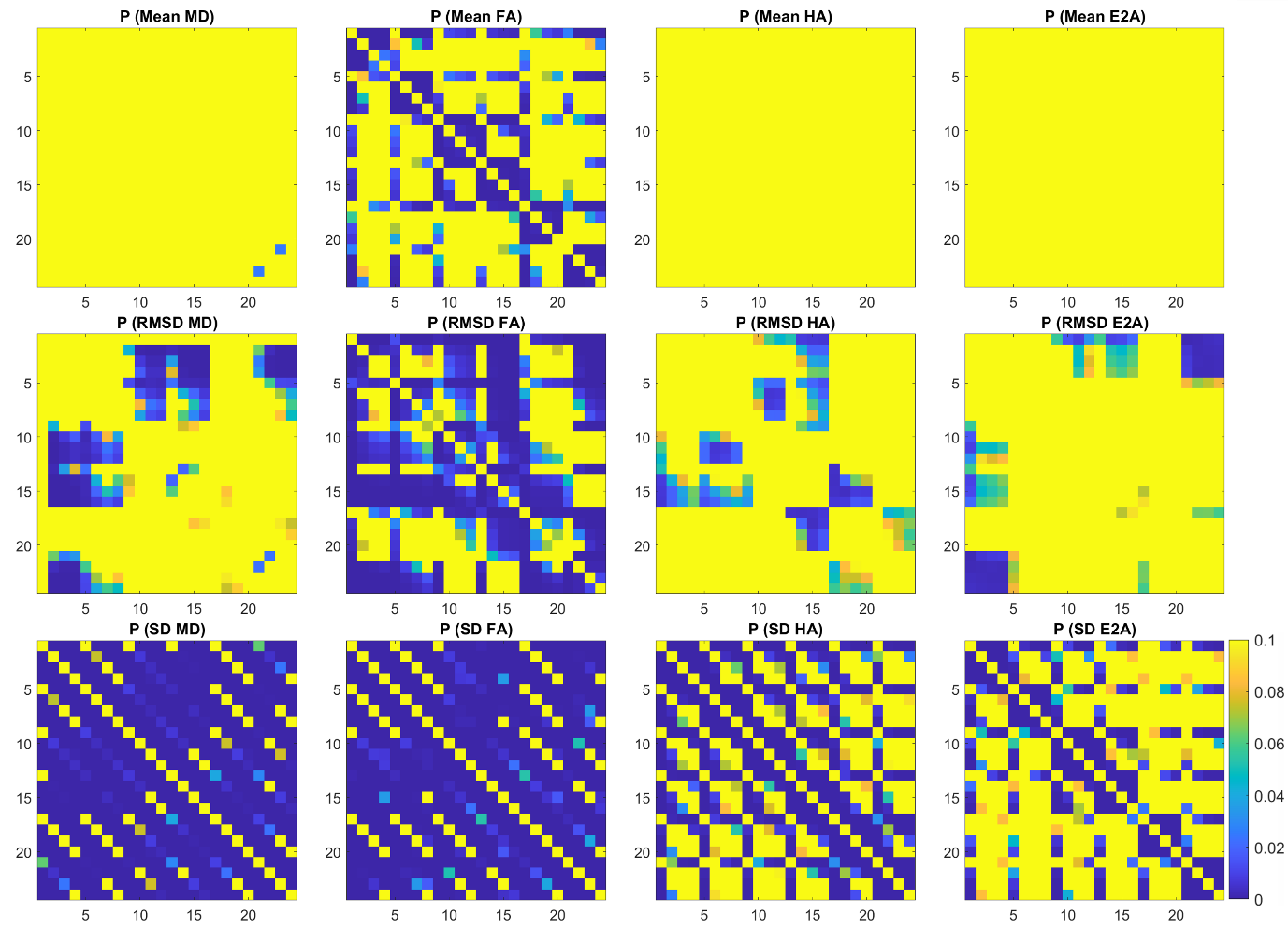

Supplementary Figure 3. P-value matrices reflecting pairwise comparisons between the 24 diffusion encoding schemes given in Supplementary Figure 1, with p < 0.05 indicating significant differences.
